# Prevalence of Strabismus in Individuals on the Autism Spectrum: A Meta-analysis

**DOI:** 10.1101/2021.07.13.21260452

**Authors:** Zachary J. Williams

**Author notes:** **Correspondence addressed to:** Zachary J. Williams, 1215 21st Avenue South, Medical Center East, Room 8310, Nashville, TN 37232. **Role of Funder/Sponsor:** No funding body or source of support had a role in the study design, data collection, analysis, or interpretation, decision to publish, or preparation of this article.

## Abstract

**Background and Objectives:** Strabismus, a misalignment of the eyes, is an important risk factor for amblyopia and visual impairment in the pediatric population. Several studies have reported an increased likelihood of strabismus in persons on the autism spectrum, but prevalence estimates in this group vary greatly.

**Methods:** We searched multiple databases to identify peer-reviewed articles published in English through November 1, 2020 that provided estimates of strabismus prevalence in autistic individuals. Prevalence estimates were synthesized using Bayesian random-effects meta-analysis, and sensitivity analysis was also performed using only the subset of studies that recruited participants from non-ophthalmologic settings and identified strabismus using structured ocular exams. Bayesian meta-regression was used to assess potential moderators of prevalence across studies.

**Results:** A total of 151 nonduplicate articles were screened, of which 22 were included in the meta-analysis (*k*=28 samples, *n*AUT=113,227). The meta-analytic point prevalence of strabismus in autistic individuals was 13.4% (95% CrI [8.3,19.4]), and sensitivity analysis produced a very similar estimate (14.0% [7.0,22.0], *n*_AUT_=581). Esotropia was the predominant subtype of strabismus reported, accounting for approximately 55% of cases. Reported prevalence rates were higher in younger samples (*BF*_10_=13.43, *R*^2^_Het_ =0.273) and samples recruited from optometry/ophthalmology clinics (*BF*_10_=11.47, *R*^2^_Het_ =0.238).

**Conclusion:** This meta-analysis found a high prevalence of strabismus in autistic individuals, with rates 3–10 times that of the general population. As untreated strabismus is a major risk factor for amblyopia in young children, these findings underscore the importance of timely screening and assessment of ocular problems in persons on the autism spectrum.

**What’s Known on This Subject:** Strabismus has been reported to be more prevalent in individuals on the autism spectrum, but estimates have been very imprecise, ranging from 3–84% across studies.

**What This Study Adds:** This study performs the first quantitative synthesis of strabismus prevalence in over 100,000 autistic individuals, generating more precise estimates of the prevalence of strabismus in the autistic population. Factors contributing to the large differences between studies are also examined using meta-regression.

## Introduction

Autism spectrum disorder (hereafter “autism”) is a lifelong neurodevelopmental condition characterized by social communication difficulties and repetitive behavior. Individuals on the autism spectrum are known to be at elevated risk for a number of co-occurring medical conditions, most notably epilepsy, gastrointestinal conditions, sleep disorders, and feeding disorders^1^. Although less well-known, autistic children and adults have high rates of ophthalmologic disorders, with rates as high as 71% across all conditions, including clinically significant refractive errors^2^.

Strabismus, a misalignment of the eyes, is a common ocular developmental disorder with onset in childhood and sequelae that frequently persist into adulthood^3^. This condition is present in 2–3% of individuals in the general population^4^ and can have profound effects on visual function and vision-related quality of life^5^. It is also a major risk factor for amblyopia, and early treatment of this condition is key to the appropriate development of binocular stereopsis. Over the last several decades, a sizable body of literature has been published on the rate of strabismus in autistic individuals, with one review citing prevalence estimates from 9–60%^6^. However, there has been no quantitative synthesis of this literature to date, substantially limiting the precision of these estimates. The current study aims to fill this gap in the literature by reporting a systematic review and meta-analysis of the prevalence of strabismus in autistic individuals, as well as the association of strabismus prevalence with a number of demographic and methodological factors.

## Methods

A systematic review and meta-analysis was conducted in accordance with PRISMA^7^ guidelines. We searched PubMed, PsycINFO, PsycARTICLES, CINAHL, ERIC, and for publications on autism that reported rates of strabismus, as defined using the following search terms: (autis^*^ OR asperger^*^ OR PDD^*^) AND (strabismus OR exotropi^*^ OR esotropi^*^ OR hypertropi^*^). Results were limited to peer-reviewed journal articles published in English before October 31, 2020. This search was supplemented with a targeted search of the archives of the *Journal of Autism and Developmental Disorders* using the keyword “strabismus” to detect articles that may have used that word only in a table. All articles included in the meta-analysis were then subjected to forward/backward citation tracing using Google Scholar to identify additional relevant studies (last search conducted November 1, 2020).

We screened titles/abstracts of all publications, using predetermined criteria to select studies for full-text review. All studies selected at this stage included conducted cross-sectional evaluations of autistic individuals that included medical exams, visual function testing, and/or review of medical records. Studies obtained by searching the *Journal of Autism and Developmental Disorders* archives were automatically all subjected to full-text review. Full-text articles were reviewed for inclusion, with the following criteria: (a) contained at least 20 individuals with diagnosed autism of any age, (b) reported data from which the proportion of autistic individuals with any form of strabismus could be derived. No authors were contacted to request unpublished data. We then extracted data from all articles according to a standardized protocol (available on request). For each included study, we extracted *n*_AUT_and *n*_Strabismus_, with multiple independent samples within an article being treated as independent studies. Potential study-level moderators were also extracted, including publication year, sample size (log transformed), proportion of females in the sample, proportion of sample with IQ/DQ<70, proportion of sample with a refractive error, mean age, method of strabismus assessment, and whether the sample was recruited from an ophthalmology/optometry clinic. Study quality was also graded using the Hoy risk of bias tool^8^.

All statistical analyses were performed in R^9^ using the *brms* package^10^. Prevalence estimates were meta-analyzed using a binomial-normal random-effects meta-analysis model with logit link function^11^, which was fit in a Bayesian framework with weakly-informative priors (Normal(0,10) prior on intercept term and half-Cauchy(0.3) prior on heterogeneity parameter τ). Heterogeneity was quantified using the *I*^*2*^ statistic and the 95% prediction interval^12^. All model parameters were summarized using the posterior median and 95% highest-density credible interval (CrI). We also conducted a sensitivity analysis, fitting the same meta-analysis model to the subset of prevalence estimates derived from studies that (a) recruited autistic participants from settings other than optometry/ophthalmology clinics, and (b) identified strabismus based on a standardized ocular exam rather than self- or proxy-report. Publication bias was assessed graphically using funnel plots.

Potential moderators of strabismus prevalence (i.e., mean age, proportion female, proportion IQ/DQ<70, proportion with refractive errors, risk of bias score, and sample recruited from optometry/ophthalmology clinic) were assessed using Bayesian meta-regression with a Normal(0,1) prior on standardized regression coefficients. Meta-regression models were compared to the baseline (intercept-only) model using a marginal likelihood-based Bayes factor (*BF*10). *BF*_10_ values >3 provided significant evidence for a moderating relationship, whereas *BF*_10_ values <1/3 provided significant evidence *against* a moderating relationship. Values between 1/3 and 3 were deemed inconclusive. Missing data were handled using 20-fold multiple imputation using the missForest algorithm^13^.

## Results

The initial database search identified 144 non-duplicate records, with an additional 7 records being identified via forward/backward citation tracing. Of these, 71 articles were subjected to full-text review, and 22 were found to meet the inclusion criteria^2,14–34^ (Figure 1). A full list of included studies can be found in Supplemental Table S1. In total, we extracted prevalence estimates from 28 independent samples, representing data from 113,227 individuals on the autism spectrum (11,697 with strabismus). Estimates of strabismus prevalence ranged from 2.5–84.3% (median 10.6%). Individuals included in the studies were predominantly school-aged children (mean age=8.56 years), 80.2% were male, and 9.7% were also diagnosed with intellectual disability, although demographics varied widely between studies. Though many studies did not report the types of strabismus observed, those that did reported a slight predominance of esotropia (*Mdn*=55.1%, *IQR* [43.6,65.6], 16 studies). Most studies showed a moderate risk of bias (*Mdn*=4, *IQR* [3.75,5]), with observed scores ranging from 1 (minimal risk of bias) to 8 (high risk of bias) on the 0–10 Hoy scale.

**Figure 1.**
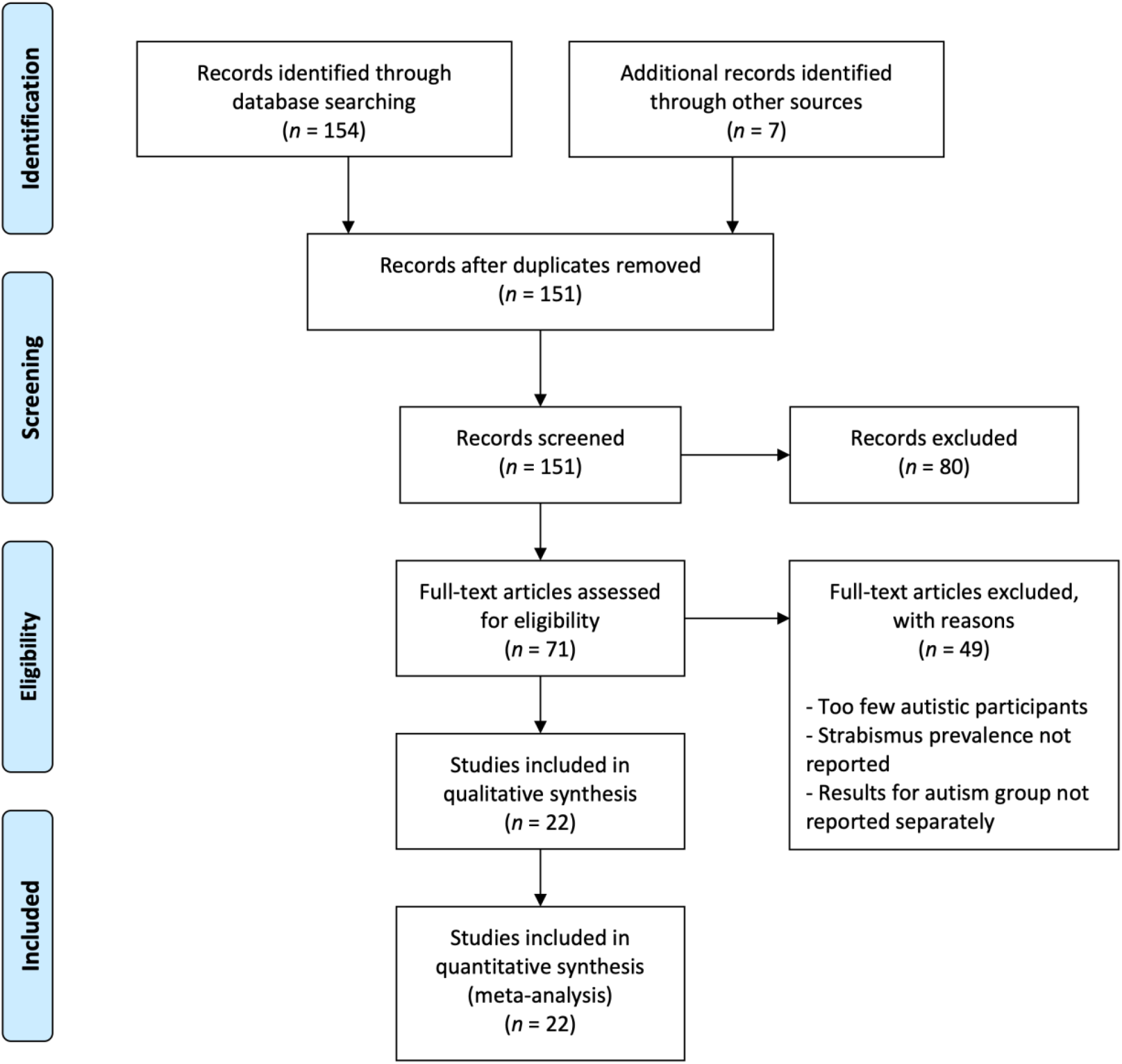
PRISMA flow diagram detailing the identification, screening and selection of studies for inclusion in the analysis. “Other sources” refers to forward- and backward-searching of included articles using Google Scholar.

Based on our meta-analytic model, the pooled point prevalence of strabismus in the autistic population was 13.4%, CrI95% [8.3,19.4] (Figure 2), with very high heterogeneity (*I*^2^=99.3% [98.9,99.7]). This value did not change appreciably when excluding the large population-based study by Chang et al., which contributed 89% of the total sample size (13.5% [8.1,19.9], *I*^2^=97.2% [95.5,98.7], *n*_AUT_=12,373). A similar estimate was also obtained when limiting our analysis to the nine studies that both recruited individuals from non-ophthalmologic settings and utilized structured eye exams to assess strabismus (14.0% [7.0,22.0], *I*^2^=79.3% [55.0,95.9], *n*_AUT_=581). The funnel plot of logit-transformed prevalence estimates (Figure 3) was largely symmetrical, suggesting minimal publication bias, although one outlying study^17^ reported a very high prevalence estimate of 84.3%.

**Figure 2.**
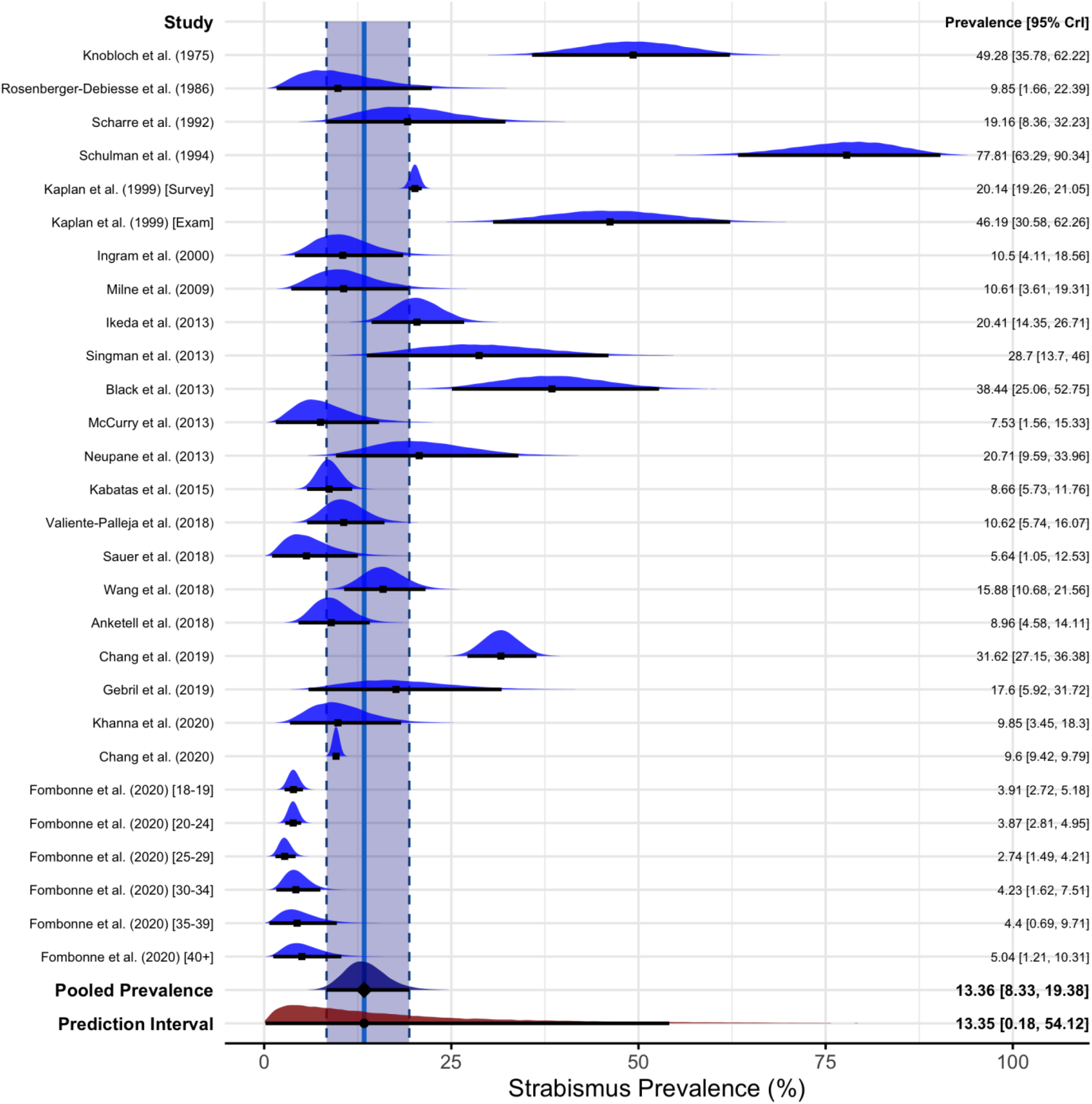
Posterior density forest plot of strabismus point prevalence in individuals on the autism spectrum. The estimated prevalence and 95% credible interval (CrI) for each study represent the posterior distribution of the prevalence in that sample, conditional on prior beliefs and the observed data. The prediction interval reports the range of prevalence values like to be observed if new studies are conducted. Density values are differentially scaled in order to visualize posterior distribution shapes for all studies. Raw prevalence values from each study can be found in Supplemental Table S1.

**Figure 3.**
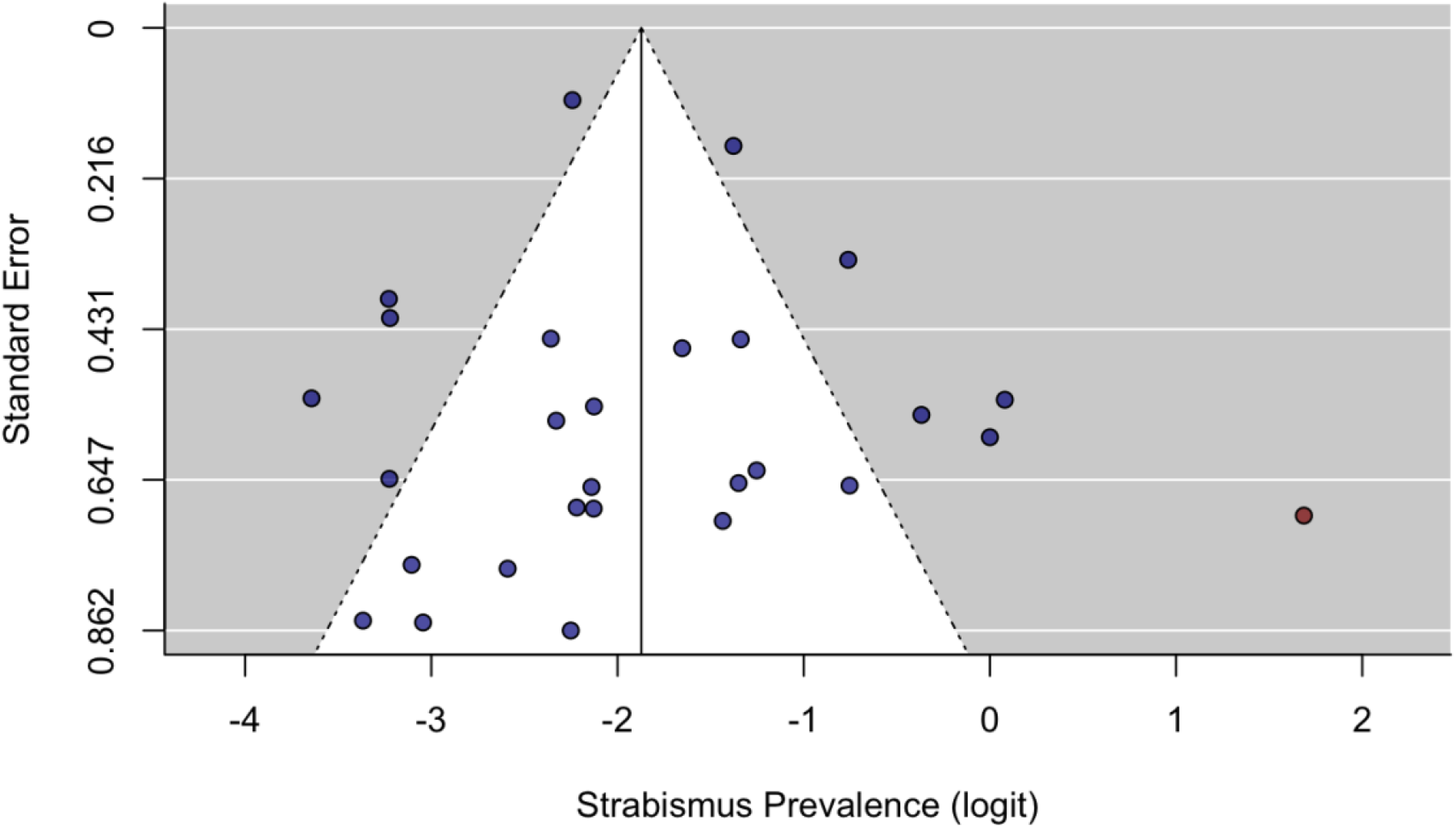
Funnel plot of strabismus prevalence estimates. With the exception of a single outlier study (Schulman, 1994, highlighted in red), the plot is relatively symmetrical, indicating little publication bias in this sample of studies.

Bayesian meta-regression indicated a that strabismus prevalence was significantly moderated by mean age, with studies of older individuals reporting lower strabismus prevalence (*BF*_10_=13.43, *R*^2^_Het_=0.273). Predicted strabismus prevalence estimates for studies with mean ages of 7.81 and 20.2 years (1 standard deviation below and above the unweighted sample mean age) were 17.4% [11.7,25.0] and 10.0% [6.4,15.1], respectively. The source of the sample was also a significant moderator (*BF*_10_=11.47, *R*^2^_Het_ =0.238), with samples recruited from optometry/ophthalmology clinics reporting substantially higher rates of strabismus (26.9% [13.3,42.3]) than samples ascertained from other sources (10.2% [6.3,15.0]). Notably, mean age continued to significantly improve the model even after controlling for sample source (*BF*10=3.28, *R*^2^_Het_ =0.360), indicating that the effect of age was not solely due to confounding by clinical samples. None of the other tested variables were significant moderators of study-level prevalence, and Bayes factors indicated substantial evidence *against* a number of these moderators (Sex ratio: *BF*_10_=0.335, proportion with intellectual disability: *BF*_10_=0.338, proportion with refractive errors: *BF*_10_=0.651, Hoy risk-of-bias score: *BF*_10_=0.244).

## Discussion

In this study, we performed the first quantitative synthesis of the literature on strabismus in the autistic population, meta-analyzing the prevalence of this condition in over 100,000 individuals on the autism spectrum. Based on our estimates, the overall point prevalence of strabismus in autistic individuals is likely between 8–20%, with similar estimates reported after eliminating a highly-influential study and limiting our analyses to studies of non-ophthalmologic samples tested using standardized ocular exams. While these estimates of co-occurring strabismus are more modest than those produced by many previous studies in autistic samples^6^, they nevertheless indicate that the prevalence of strabismus in autism is 3–10 times that of the general population. Pediatricians and other clinicians examining autistic individuals should be aware of this increased risk for strabismus and consider referring patients to pediatric ophthalmologists for more comprehensive examinations of ocular pathology.

Strabismus prevalence across studies was moderated both by sample age (lower prevalence in older samples) and sample source (higher prevalence in samples recruited from optometry/ophthalmology clinics), although there was evidence against moderation by sex, proportion of individuals with intellectual disability, and risk-of-bias scores. Perhaps unsurprisingly, samples recruited from ophthalmologic settings had nearly three times the prevalence of strabismus (27%) than samples recruited from other sources (10.2%), confirming prior suspicions that prevalence estimates from many prior studies in autistic individuals are inflated due to biased recruitment^6^. Studies of older individuals were also likely to report lower strabismus prevalence, potentially due to either medical interventions or spontaneous remission of strabismus with increasing age. However, longitudinal studies of strabismus in autism are necessary to confidently determine the clinical course of strabismus and associated impairments in this population. Notably, a large portion of heterogeneity (64%) remained unexplained after accounting for these two significant moderators, indicating that vast majority of between-study heterogeneity is not attributable to sample demographics or easily quantified study-level variables.

Importantly, this study is not without its limitations. One major limitation was that definitions of strabismus were not standardized across studies, with questionnaires, exam findings, and ICD codes all combined to produce the overall prevalence estimates. However, in our sensitivity analyses, we included studies that only assessed strabismus using ocular exam techniques, which, while somewhat different between studies, were substantially less heterogeneous yet produced a very similar prevalence estimate. Additionally, only one of the included studies^33^ was truly population-based, and the remainder utilized samples of convenience from community, clinical, or registry sources. Their representativeness of the overall autistic population may vary drastically, biasing the included prevalence estimates. Population-based epidemiologic studies such as large-scale medical record reviews in Nordic countries may further increase the precision of these estimates. As an additional limitation, the reviewed studies did not generally provide information about the degree of strabismus or the rates of amblyopia or other sequelae directly related to strabismus in this population. Future epidemiologic studies of strabismus in autism should specifically attempt to investigate the severity and impact of strabismus in the autistic population, thereby addressing this gap in the literature. Lastly, while our meta-regression failed to find associations between strabismus and factors such as refractive error or intellectual disability, such analyses are substantially underpowered compared to individual participant analyses and should be confirmed with large-sample individual-participant analyses.

## Conclusions

In conclusion, our meta-analysis suggests that strabismus is substantially more common in individuals on the autism spectrum than the general population, putting autistic persons at increased risk for amblyopia and other serious sequelae of this condition. The findings of this study highlight the need to screen individuals diagnosed with autism for ocular problems early in life, as early correction of strabismus may significantly decrease the incidence of amblyopia in these individuals and improve overall quality of life^35^. Further studies of strabismus outcomes in autistic children and adults are necessary to determine whether standard-of-care strabismus treatments remain the most appropriate clinical interventions for individuals in this population.

## Data Availability

Original data used in the meta-analysis can be found in the primary articles, as well as supplemental table 1.

## Acknowledgements

The author would like to thank the research teams who conducted all primary studies summarized herein, as well as all of the autistic individuals and their families who have contributed to this body of research.

## Contributors’ Statement

**Zachary J. Williams:** Mr. Williams conceptualized and designed the study, performed the literature search and article selection, conducted all statistical analyses, and drafted the manuscript. He approves the final manuscript as submitted and agrees to be accountable for all aspects of the work.

## Abbreviations

*BF*_10_: Bayes factor comparing meta-regression model to baseline model
CrI: Highest-density credible interval
DQ: Developmental quotient
IQ: Intelligence quotient
*IQR*: Interquartile range
*Mdn*: Median
*n*_AUT_: Number of individuals diagnosed with autism in a study
*n*_Strabismus_: Number of autistic individuals diagnosed with strabismus in a study
*R*^2^_Het_: Proportion of heterogeneity explained by moderator

---

**Supplemental Table S1.** Samples included in meta-analysis of strabismus prevalence

| Sample | $n_{\text{AUT}}$ | $n_{\text{Strabismus}}$ | Prevalence (%) | Esotropia (%) | Opt Exam | Female (%) | ID/DD (%) | Refract (%) | Age (Years) | Country | Opt Cohort | ROB (0–10) |
| --- | --- | --- | --- | --- | --- | --- | --- | --- | --- | --- | --- | --- |
| Knobloch et al. (1975) | 50 | 26 | 52.0 | — | FALSE | — | 80.0 | — | 1.5 | USA | FALSE | 8 |
| Rosenberger-Debiesse et al. (1986) | 21 | 2 | 9.5 | — | FALSE | 42.9 | — | — | 11.7 | France | FALSE | 8 |
| Scharre et al. (1992) | 34 | 7 | 20.6 | 14.3 | TRUE | 5.9 | 44.1 | — | 7.5 | USA | FALSE | 3 |
| Schulman et al. (1994) | 32 | 27 | 84.4 | 55.6 | TRUE | 25.0 | — | 37.0 | — | USA | TRUE | 5 |
| Kaplan et al. (1999) [Survey] | 7640 | 1539 | 20.1 | — | FALSE | 21.4 | — | — | 7.8 | USA | FALSE | 6 |
| Kaplan et al. (1999) [Exam] | 34 | 17 | 50.0 | 35.0 | TRUE | — | — | — | 13.4 | Canada | FALSE | 3 |
| Ingram et al. (2000) | 57 | 6 | 10.5 | 33.3 | FALSE | 19.3 | 75.4 | — | — | USA | FALSE | 8 |
| Milne et al. (2009) | 47 | 5 | 10.6 | 60.0 | FALSE | 13.7 | 23.4 | 21.6 | 12.1 | UK | FALSE | 4 |
| Ikeda et al. (2013) | 154 | 32 | 20.8 | 62.5 | TRUE | 20.8 | 18.0 | 28.6 | 3.3 | USA | TRUE | 3 |
| Singman et al. (2013) | 25 | 8 | 32.0 | 75.0 | TRUE | — | — | — | 6.0 | USA | TRUE | 4 |
| Black et al. (2013) | 44 | 18 | 40.9 | 50.0 | TRUE | 25.0 | — | 27.3 | 11.0 | USA | TRUE | 4 |
| McCurry et al. (2013) | 43 | 3 | 7.0 | — | TRUE | 28.1 | — | 44.2 | 9.3 | Peru | FALSE | 4 |
| Neupane et al. (2013) | 36 | 8 | 22.2 | 25.0 | TRUE | 25.0 | — | 58.3 | 9.0 | Nepal | FALSE | 4 |
| Kabatas et al. (2015) | 324 | 28 | 8.6 | 46.4 | TRUE | 17.6 | 4.9 | 22.5 | 5.0 | Turkey | TRUE | 4 |
| Valiente-Palleja et al. (2018) | 122 | 13 | 10.7 | — | FALSE | 34.4 | 100.0 | — | 40.7 | Spain | FALSE | 8 |
| Sauer et al. (2018) | 44 | 2 | 4.5 | 100 | TRUE | 11.4 | 87.2 | 15.9 | 2.2 | Peru | FALSE | 3 |
| Wang et al. (2018) | 168 | 27 | 16.1 | 88.9 | TRUE | 14.9 | — | 22.0 | 5.0 | China | FALSE | 4 |
| Anketell et al. (2018)<sup>a</sup> | 124 | 11 | 8.9 | 54.5 | TRUE | 18.8 | — | 86.7 | 10.9 | UK | FALSE | 3 |
| Chang et al. (2019) | 380 | 121 | 31.8 | 49.6 | TRUE | 23.4 | 15.8 | 41.8 | 5.5 | USA | TRUE | 4 |
| Gebril et al. (2019) | 26 | 5 | 19.2 | 100 | TRUE | 46.2 | — | 80.8 | 6.4 | Libya | TRUE | 5 |
| Khan— et al. (2020) | 51 | 5 | 9.8 | 60 | TRUE | 15.7 | 61.8 | 35.3 | 5.6 | France | FALSE | 1 |
| Chang et al. (2020) | 100854 | 9682 | 9.6 | — | FALSE | 19.6 | 8.3 | — | 8.2 | USA | FALSE | 3 |
| Fombonne et al. (2020) [18-19] | 913 | 35 | 3.8 | — | FALSE | 23.0 | 37.5 | — | 18.5 | USA | FALSE | 4 |
| Fombonne et al. (2020) [20-24] | 1207 | 46 | 3.8 | — | FALSE | 20.6 | 47.0 | — | 22.0 | USA | FALSE | 4 |
| Fombonne et al. (2020) [25-29] | 510 | 13 | 2.5 | — | FALSE | 18.6 | 55.3 | — | 27.0 | USA | FALSE | 5 |
| Fombonne et al. (2020) [30-34] | 157 | 6 | 3.8 | — | FALSE | 22.9 | 65.0 | — | 32.0 | USA | FALSE | 5 |
| Fombonne et al. (2020) [35-39] | 60 | 2 | 3.3 | — | FALSE | 21.7 | 70.0 | — | 37.0 | USA | FALSE | 5 |
| Fombonne et al. (2020) [40+] | 70 | 3 | 4.3 | — | FALSE | 18.6 | 62.9 | — | 45.0 | USA | FALSE | 5 |

*Note.* Fields marked with “—” were not reported in the study. $n_{\text{AUT}}$ = number of autistic individuals in the study cohort; $n_{\text{Strabismus}}$ = number of autistic individuals with identified strabismus; Opt Exam = ophthalmologic; ID/DD = proportion of individuals in the sample with intellectual disability or developmental delay; Refract = proportion of individuals in the sample with refractive errors; ROB = Hoy risk-of-bias score for prevalence studies (ranges from 0 [no bias] to 10 [extreme bias]).
<sup>a</sup> Data from this study is drawn from the linked paper, as well as two other reports on the same data (Anketell et al., 2016, *Ophthalmic Physiol Op*; Little et al., 2018 *Clin Exp Optom*)

